# Discrepancy between knowledge and practice of self-medication among adults in Dhaka, Bangladesh in 2023

**DOI:** 10.1101/2025.06.29.25330523

**Authors:** Salma Sultana, Immamul Muntasir, Mohammad Asraful Alam

## Abstract

**Introduction:** Self-medication, while convenient for managing minor illnesses, poses significant public health risks when practiced without adequate knowledge or oversight. In Bangladesh, especially in urban areas, self-medication is widespread due to limited healthcare access and weak pharmaceutical regulation. This study aims to assess the knowledge, attitudes, and practices of self-medication among urban adult residents in Dhaka, Bangladesh.

**Methods:** A cross-sectional descriptive study was conducted from January to December 2023 in Badda, Dhaka. A total of 361 adults who attempted to purchase medications without a prescription were interviewed using a semi-structured questionnaire. Data were analyzed using descriptive statistics and chi-square tests for associations.

**Results:** Among participants, 54.8% reported self-medicating within the past three months. Commonly treated ailments included cough, headache, and acidity. Analgesics, antiulcerants, and antibiotics were frequently used. While 64.8% claimed knowledge of the medicines used, many modified dosages or discontinued treatment early. Notably, 41.3% experienced side effects. Although 85.3% believed self-medication was harmful, the practice persisted due to convenience and cost barriers. Higher education was significantly associated with greater awareness of medication side effects (p = 0.002).

**Conclusion:** Despite general awareness of risks, self-medication remains prevalent and often unsafe among urban adults in Dhaka. Strengthening public education, regulatory enforcement, and healthcare access is essential to promote safer self-care behaviors.

## Introduction

Self-medication is defined as obtaining and using medications without a physician’s advice[1]. It is an integral part of self-care, encompassing the use of over-the-counter (OTC) drugs and home remedies for minor conditions[2]. The World Health Organization notes that responsible self-medication can relieve the burden on healthcare systems and provide prompt relief for self-diagnosed symptoms[2]. For example, individuals often treat headaches, colds, or mild gastrointestinal complaints with OTC medicines, bypassing formal medical consultations[2].

However, self-medication carries potential risks, especially when medications are used irrationally or without adequate knowledge. Unsupervised drug use can lead to incorrect dosing, adverse reactions, dangerous drug interactions, masking of serious diseases, and the development of antimicrobial resistance[1,3,4]. In particular, the improper use of antibiotics for self-treatment is a serious global concern[3,5]. While self-medication can conserve medical resources when done responsibly, its widespread practice in many countries has highlighted the need for improved awareness and regulation[3,5].

In Bangladesh, systemic factors encourage self-medication. Healthcare workforce shortages and limited infrastructure (including only about 13 health workers per 10,000 population) make access to doctors difficult, especially for low-income or rural populations[4,6]. Low public health spending and lack of insurance further limit formal care. Moreover, weak drug regulation and prevalent over-the-counter sales of prescription medicines exacerbate the problem. Previous studies in Bangladesh have reported very high self-medication rates (16–81% in urban areas) with common reasons including perceived mild illness and cost or time barriers to visiting a doctor[3,4,7–12].

Given these concerns, it is important to understand how urban residents in Dhaka – a densely populated, resource-constrained setting – approach self-medication. This study aimed to assess the knowledge, attitudes, and practices related to self-medication among adult pharmacy customers in Dhaka who obtained medicines without prescriptions. Identifying their behaviors and perceptions can inform public health strategies to promote safer medication practices.

## Materials and Methods

### Study Design and Setting

This descriptive cross-sectional study was conducted from January to December 2023 in the Badda area of Dhaka North City Corporation, Bangladesh. Badda is a densely populated urban neighborhood comprising residential, commercial, and informal settlements. The area was selected due to its demographic diversity and high reliance on community pharmacies for healthcare needs, making it a representative setting for studying real-world self-medication practices in an urban Bangladeshi context.

### Study Population and Inclusion Criteria

The target population included adults aged 18 years and above who resided in Badda and had attempted to purchase or recently purchased medications without a valid medical prescription. Inclusion required that participants be capable of providing informed consent and willing to participate in a face-to-face interview.

### Sample Size Calculation

The sample size was calculated using the standard formula for estimating proportions in cross-sectional studies. With a 95% confidence level, a 5% margin of error, and an expected prevalence of 48.5% (based on reported urban self-medication rates ranging from 16% to 81%), the minimum required sample size was determined to be 384. A total of 361 participants were enrolled, which was considered adequate given logistical constraints and the descriptive nature of the study.

### Sampling Strategy

A convenience sampling method was used. Community pharmacies in Badda were selected based on patient volume, geographic accessibility, and willingness of staff to participate. Pharmacies with high customer flow in residential and mixed-use areas were prioritized to ensure that data reflected typical patterns of urban self-medication behavior. This approach allowed efficient recruitment while maintaining ecological validity for the urban population.

### Data Collection Tool and Method

Data were collected through face-to-face interviews using a semi-structured questionnaire developed through literature review and expert consultation. The instrument was pre-tested with 20 individuals in a neighboring area to ensure clarity and reliability. It included questions on socio-demographic characteristics, recent illness episodes, self-medication behavior, drug types used, reasons for self-medication, knowledge of side effects, and attitudes toward the practice.

Trained data collectors conducted the interviews in private areas of participating pharmacies. Written informed consent was obtained from all participants prior to data collection.

### Ethical Considerations

The study received ethical approval from the Ethical Review Committee of the National Institute of Preventive and Social Medicine (NIPSOM), Dhaka, Bangladesh. Participation was voluntary, and all data were collected anonymously to ensure confidentiality.

### Data Analysis

Data were entered and cleaned in IBM SPSS Statistics 25.0. Descriptive statistics (frequencies and percentages) were used to summarize categorical variables. The chi-square test and Fisher’s exact test were used to examine associations between demographic variables and knowledge or behavior related to self-medication. A p-value <0.05 was considered statistically significant.

**Map:**
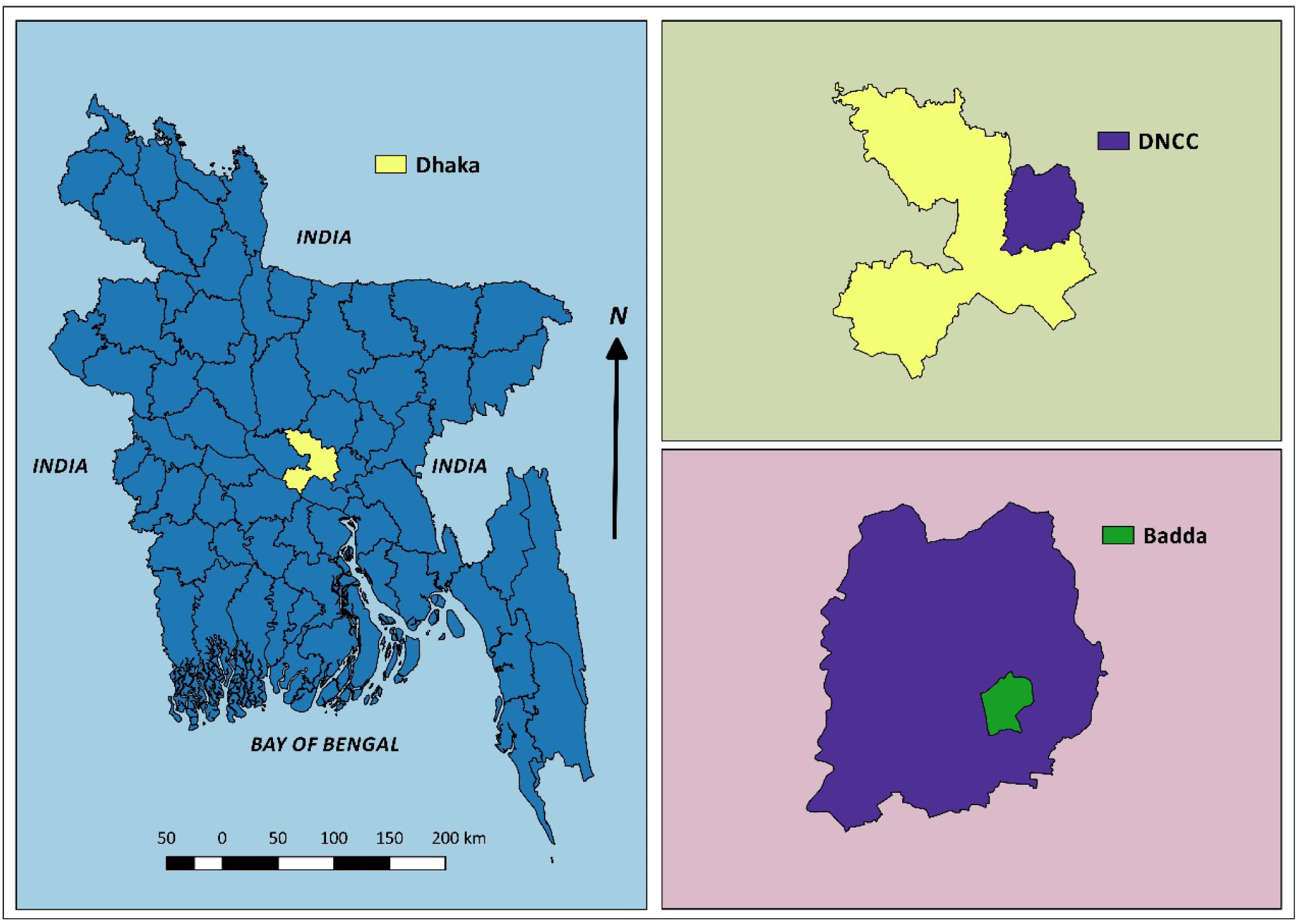
Area of the study on self-medication (SM) knowledge, practice and attitudes of urban residents of a selected neighborhood of Dhaka, Bangladesh in 2023.

## Results

A total of 361 adults participated. As shown in **Table 1**, most participants were male (74.5%). The largest age group was 18–27 years (40.2%), followed by 28–37 years (29.9%). Nearly all (91.1%) were Muslim, and 59.0% had ever been married. About one-third (31.6%) had a graduate degree, and 20.2% had completed higher secondary education. The most common occupations were service holders (28.5%) and students (26.9%). Most lived in nuclear families (64.3%), and about 62.6% reported 4–6 family members. Monthly family income was often in the mid-range: the largest group (37.1%) earned between 15,001 and 30,000 Taka.

**Table 1:** Socio-demographic characteristics of participants (n=361)

| Characteristics | Category | n | % |
| --- | --- | --- | --- |
| Age group (years) | 18–27 | 145 | 40.2 |
|  | 28–37 | 108 | 29.9 |
|  | 38–47 | 52 | 14.4 |
|  | 48–57 | 31 | 8.6 |
|  | ≥58 | 25 | 6.9 |
| Gender | Male | 269 | 74.5 |
|  | Female | 92 | 25.5 |
| Education | Graduate degree | 114 | 31.6 |
|  | Higher secondary (A-level) | 73 | 20.2 |
|  | Secondary | 51 | 14.1 |
|  | Others/less | — | — |
| Occupation | Service holder | 103 | 28.5 |
|  | Student | 97 | 26.9 |
|  | Business | 58 | 16.1 |
|  | Housewife/Other | 103 | 28.5 |
| Family type | Nuclear | 232 | 64.3 |
|  | Joint | 129 | 35.7 |
| Family size | ≤3 members | 87 | 24.1 |
|  | 4–6 members | 226 | 62.6 |
|  | ≥7 members | 48 | 13.3 |
| Monthly income (Taka) | ≤15,000 | 120 | 33.2 |
|  | 15,001–30,000 | 134 | 37.1 |
|  | 30,001–45,000 | 66 | 18.3 |
|  | >45,000 | 41 | 11.4 |

Within the three months prior to the survey, 330 participants (91%) reported experiencing at least one illness, and 198 of these (54.8%) admitted to self-medicating for their condition (**Table 2**). Among those who self-medicated, the most common motivations were: belief that a doctor’s visit was unnecessary for minor illnesses (32.8%), having a previous prescription on hand (26.3%), pharmacist advice (11.6%), and high doctor’s fees (10.6%). Other reasons (such as long waiting times or confidence in self-diagnosis) were less frequently mentioned. For the 163 ill participants who did *not* self-medicate, the leading concerns were fear of using the wrong drug (30.7%) and lack of knowledge about the medication (25.8%). A smaller proportion cited fear of adverse effects or drug interactions. These findings indicate that safety concerns deterred nearly one-third of patients from self-medicating.

**Table 2:** Distribution by disease occurrence and practice of self-medication in prior three months.

| Characteristics | Categories | n | Percent (%) |
| --- | --- | --- | --- |
| Disease occurrence in the prior three months | Yes | 330 | 91 |
|  | No | 31 | 9 |
| Incidence of taking self-medication in last three months | No | 163 | 45.2 |
|  | Yes | 198 | 54.8 |
| Reasons for not taking self-medication (n = 163) | Lack of knowledge about medicine | 42 | 25.8 |
|  | Risk of adverse effect | 24 | 14.7 |
|  | Risk of using wrong drugs | 50 | 30.7 |
|  | Risk of misdiagnosing | 23 | 14.1 |
|  | Risk of using drug wrongly | 13 | 8.0 |
|  | Others | 11 | 6.7 |
| Reasons for taking self-medication in the last three months (n = 198) | Time saving | 9 | 4.5 |
|  | High fees of doctors | 21 | 10.6 |
|  | Having old prescription | 52 | 26.3 |
|  | Long waiting time | 10 | 5.1 |
|  | No need to visit doctor for minor illness | 65 | 32.8 |
|  | Pharmacist's advice | 23 | 11.6 |
|  | Confidence on their knowledge about medicines | 9 | 4.5 |
|  | Other reasons | 9 | 4.5 |

Among the types of ailments treated by self-medication, cough (41.3%), headache/migraine (34.1%), fever (31.6%), body ache (28.3%), and acidity (26.0%) were most prevalent (**Table 3**). Eye infections, ulcers, allergies, vomiting, and skin diseases were reported less often. Nearly all participants (86.1%) used allopathic (Western) medications; only 6.1% reported using herbal or Ayurvedic remedies, and the remainder used home remedies. The most commonly used drug classes were analgesics (47.9%) and antiulcerants (47.1%), followed by antibiotics (30.5%), antihistamines (13.3%), and antipyretics.

**Table 3:** Conditions treated and medications used for self-medication (n=198)

| Characteristics | Categories | n | Percent (%) |
| --- | --- | --- | --- |
| Diseases for which the respondents took self-medication* | Headache / migraine | 123 | 34.1 |
|  | Eye infection | 19 | 5.3 |
|  | Cough | 149 | 41.3 |
|  | Body ache | 102 | 28.3 |
|  | Mouth ulcer | 26 | 7.2 |
|  | Allergy | 48 | 13.3 |
|  | Acidity | 94 | 26.0 |
|  | Vomiting | 87 | 24.1 |
|  | Fever | 114 | 31.6 |
|  | Skin disease | 41 | 11.4 |
|  | Diarrhea | 43 | 11.9 |
|  | Insomnia | 33 | 9.1 |
| Form of medicine preferred | Ayurvedic | 22 | 6.1 |
|  | Allopathic | 311 | 86.1 |
|  | Homeopathic | 14 | 3.9 |
|  | Unani | 14 | 3.9 |
| Types of medicine taken* | Antibiotic | 110 | 30.5 |
|  | Analgesic | 173 | 47.9 |
|  | Antiulcerant | 170 | 47.1 |
|  | Antihistamine | 128 | 35.5 |
|  | Cough syrup | 99 | 27.4 |
|  | Vitamins | 105 | 29.1 |
|  | Laxatives | 24 | 6.6 |
|  | Antiemetic | 79 | 21.9 |
|  | Antipyretic | 122 | 33.8 |
| Knowledge about the medicines | Yes | 234 | 64.8 |
|  | No | 127 | 35.2 |
| Frequency of self-medication | Regularly | 74 | 20.5 |
|  | Irregularly | 173 | 47.9 |
|  | Rarely | 114 | 31.6 |
| Reason for selecting the particular medicine* | Recommended by pharmacist | 120 | 33.2 |
|  | Old prescription of doctor | 156 | 43.2 |
|  | Used by peers/friend/family | 11 | 3.0 |
|  | Respondent's previous experience | 67 | 18.6 |
|  | Other reason | 7 | 2.0 |
| Source of obtaining drug for self-medication | Pharmacy | 354 | 98.1 |
|  | Others | 7 | 1.9 |

\*Multiple response

About 64.8% of self-medicators stated they had knowledge about the medicines they took. However, usage patterns varied: 47.9% reported taking medications on an irregular schedule (e.g. not following a fixed dosing regimen), 36.9% sometimes followed a regular schedule, and 15.1% always took them as directed. The main reasons for choosing specific medicines were having an old prescription (43.2%) and pharmacist recommendation (33.2%); only 9.4% selected drugs based on confidence in their own knowledge. Almost all (98.1%) obtained their medications directly from pharmacies; very few relied on leftover household drugs or borrowed from others. (**Table 3**)

We also examined practices related to prescription use and adherence (**Table 4**). About half of participants (49.6%) said they sometimes checked a previous prescription before using the same medication again, while 21.3% always did and 29.1% never did. In terms of understanding instructions, only 20.5% fully understood the directions on a prescription, 51.0% partially understood them, and 28.5% did not understand at all. Similarly, only 14.1% always read the package instructions on medicines, 44.0% did so sometimes, and 41.9% rarely or never.

**Table 4:**
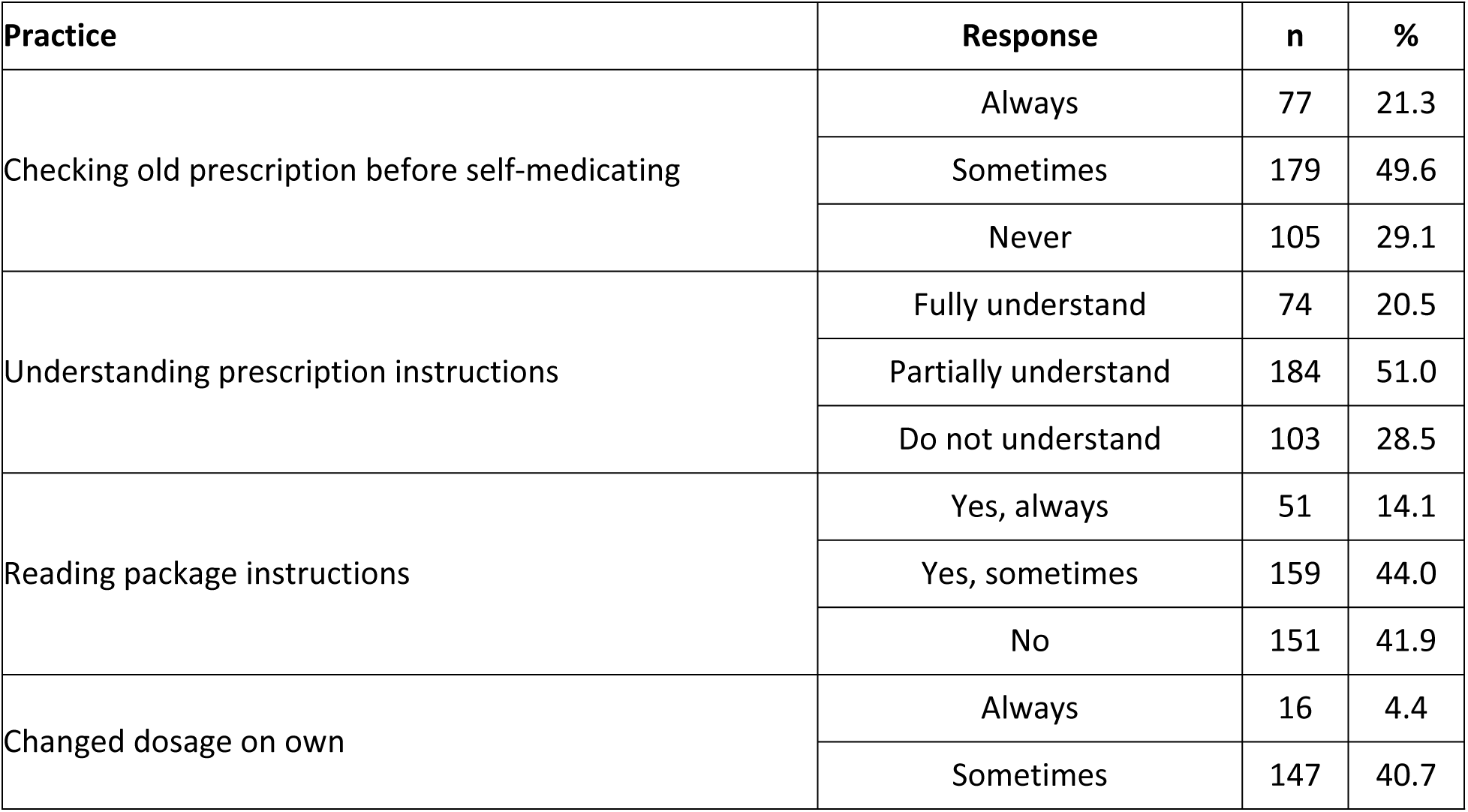
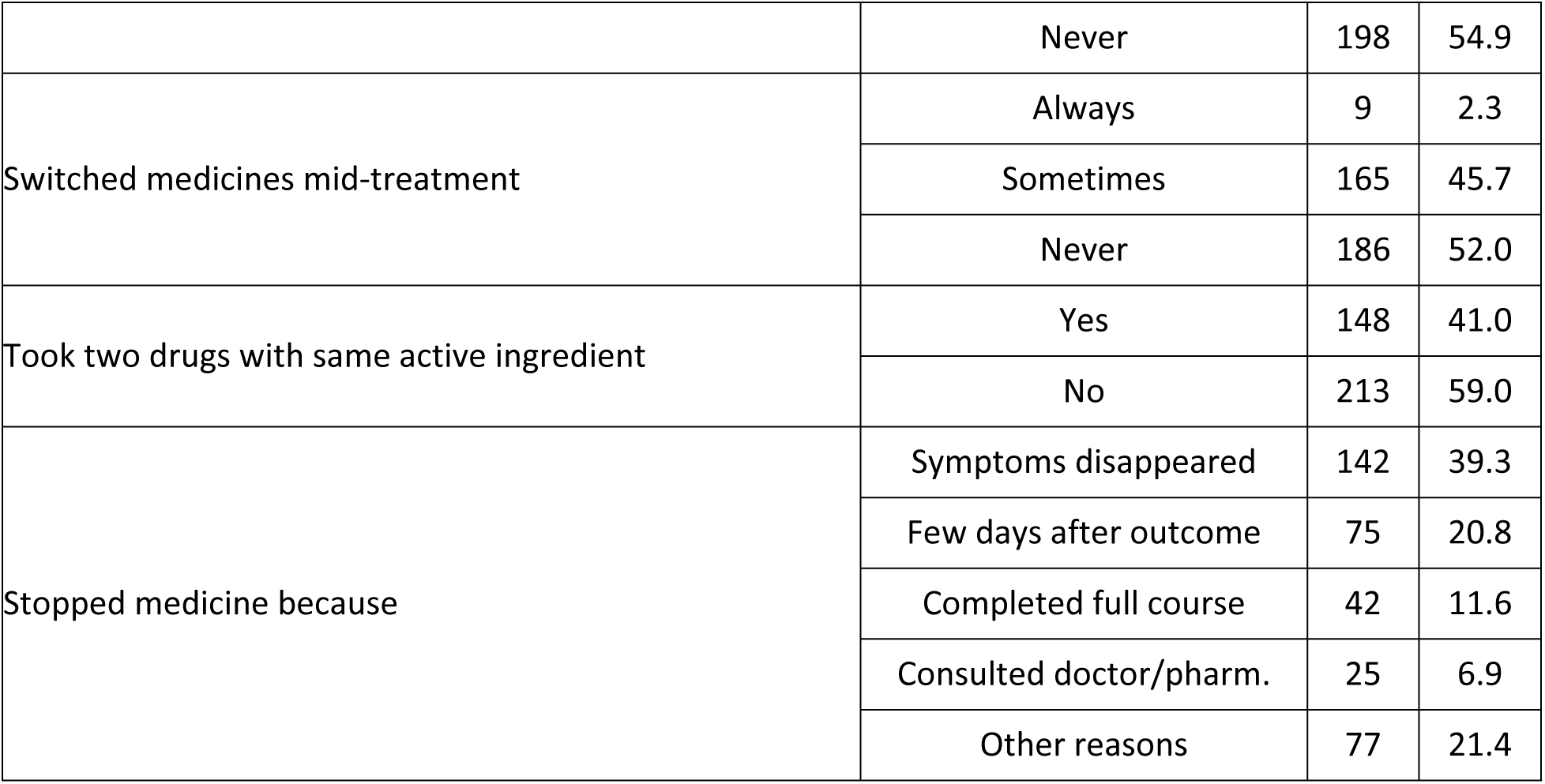
Practices related to prescription use and medication adherence (n=361)

| Practice | Response | n | % |
| --- | --- | --- | --- |
| Checking old prescription before self-medicating | Always | 77 | 21.3 |
|  | Sometimes | 179 | 49.6 |
|  | Never | 105 | 29.1 |
| Understanding prescription instructions | Fully understand | 74 | 20.5 |
|  | Partially understand | 184 | 51.0 |
|  | Do not understand | 103 | 28.5 |
| Reading package instructions | Yes, always | 51 | 14.1 |
|  | Yes, sometimes | 159 | 44.0 |
|  | No | 151 | 41.9 |
| Changed dosage on own | Always | 16 | 4.4 |
|  | Sometimes | 147 | 40.7 |
|  | Never | 198 | 54.9 |
| Switched medicines mid-treatment | Always | 9 | 2.3 |
|  | Sometimes | 165 | 45.7 |
|  | Never | 186 | 52.0 |
| Took two drugs with same active ingredient | Yes | 148 | 41.0 |
|  | No | 213 | 59.0 |
| Stopped medicine because | Symptoms disappeared | 142 | 39.3 |
|  | Few days after outcome | 75 | 20.8 |
|  | Completed full course | 42 | 11.6 |
|  | Consulted doctor/pharm. | 25 | 6.9 |
|  | Other reasons | 77 | 21.4 |

A substantial fraction reported modifying their treatment: 40.7% occasionally changed the dosage of a medication on their own (e.g., taking more or less than directed), and 45.7% sometimes switched to a different medicine during an illness course. Notably, 41.0% admitted to taking two different drugs that contained the same active ingredient at the same time. The most common reason given for stopping a medication was symptom resolution (39.3% stopped once they felt better). Only 11.6% of self-medicators completed the full recommended course of treatment, and just 6.9% ceased treatment only after consulting a doctor or pharmacist.

Regarding side effect awareness and response, 271 participants (75.1%) reported that they knew about the potential side effects of medicines. The most commonly recognized risks (multiple answers allowed) were kidney damage (44.8%), liver damage (28.7%), and weakened immune function (29.1%). Despite this knowledge, 75 out of the 198 self-medicators (41.3%) experienced some side effect from their medication. When side effects occurred, 40.3% went to a physician’s private chamber, 26.8% consulted a pharmacist, 20.1% simply stopped the medication, and only 11.4% sought care at a formal health facility.

Finally, participants’ perceptions of self-medication were predominantly negative (**Table 5**). A large majority (85.3%) believed that self-medication is harmful. Most participants (76.7%) regarded self-medication as unacceptable for one’s own healthcare, with only 6.9% considering it a good or acceptable practice. Likewise, 70.1% said they would not recommend self-medication to others. About half (52.9%) were aware that there are legal restrictions on dispensing prescription drugs, but only 11.1% had personally encountered any legal consequences. Overall, 89.5% of participants agreed that self-medication practices should be stopped or discouraged.

**Table 5:** Perceptions of self-medication (n=361)

| **Perception** | **Response** | **n** | **%** |
| --- | --- | --- | --- |
| Attitude towards self-medication | Not acceptable practice | 277 | 76.7 |
|  | Acceptable practice | 59 | 16.3 |
|  | Good practice | 25 | 6.9 |
| Confidence in self-treatment | No, I cannot | 210 | 58.2 |
|  | Not sure | 116 | 32.1 |
|  | Yes, I can | 35 | 9.7 |
| Belief that self-medication is harmful | Yes | 308 | 85.3 |
|  | No | 53 | 14.7 |
| Recommend self-medication to others | No | 253 | 70.1 |
|  | Yes | 108 | 29.9 |
| Aware of legal barriers | Yes | 191 | 52.9 |
|  | No | 170 | 47.1 |
| Experienced legal issues | Yes | 40 | 11.1 |
|  | No | 321 | 88.9 |

## Discussion

This study found that self-medication is widely practiced among urban adults in Dhaka, often for convenience and cost-saving. The majority of users were young, educated males, which aligns with other research in South Asia suggesting that higher education and easy information access can promote self-treatment behaviors. The overall self-medication rate (54.8% of those with an illness) falls within the high range reported in similar settings. For example, studies in Bangladesh and India have documented prevalent self-medication for minor complaints such as headaches and colds. The main reasons identified here – perceiving ailments as minor and relying on past prescriptions – echo findings from previous studies in Bangladesh and neighboring countries.

In our study, respiratory and general symptoms were most commonly self-treated: cough (41.3%), headache (34.1%), fever (31.6%), body ache (28.3%), and acidity (26%). These patterns are similar to other reports from the region, where such non-serious conditions frequently lead individuals to self-medicate[1,9,10,13–15]. As expected, allopathic drugs dominated the practice (86.1%), with traditional remedies used by only a small fraction. Analgesics and antiulcerants were the top classes of medications (each used by about 47% of participants), followed by antibiotics (30.5%). The high use of antibiotics without prescription is concerning given the global threat of resistance. This rate is comparable to other Bangladeshi surveys, though some have reported even higher antibiotic self-use[3,5].

We observed that many participants did not follow recommended medication regimens. Nearly half took drugs irregularly or altered dosages on their own. For instance, 40.7% occasionally changed doses and 45.7% switched to different medicines mid-course. Very few completed full treatment courses. This is consistent with reports from other countries: one Chinese study found that over half of self-medicated students altered antibiotic use and dosages[16]. Importantly, 41.0% of our participants admitted to taking two medications with the same active ingredient at once, risking overdose or increased side effects. These behaviors highlight the potential dangers of unsupervised medication use.

Despite this, many participants were knowledgeable about risks. Three-quarters (75.1%) knew that medicines can cause side effects, and nearly half could identify serious complications like kidney or liver damage. This awareness appears higher than in some other studies (for example, some Indian studies reported very low awareness of drug side effects)[17]. Nonetheless, 41.3% of our self-medicating subjects experienced adverse effects, and most sought help appropriately (40.3% saw a physician). This suggests that personal experience may reinforce caution.

Notably, attitudes toward self-medication were predominantly negative. The vast majority (85.3%) considered self-medication harmful, and almost 90% agreed that the practice should cease. Most did not see self-medication as acceptable for personal use and would not recommend it to others. These findings indicate a significant level of insight into the risks, despite the continued practice. Such negative perceptions are encouraging, as they suggest that public health messages could build on existing concerns. Only a small minority was confident in their own ability to self-treat serious conditions (9.7%), further underscoring skepticism about unsupervised care. A significant association was found between educational qualification and awareness of medication side effects (p = 0.002), indicating that participants with higher education levels were more likely to be informed about the potential risks of self-medication.

This study has limitations. The convenience sample from one area of Dhaka may not represent all urban populations. Self-reported practices may be subject to recall bias. Additionally, we focused on adult pharmacy customers and did not include individuals who may self-medicate from home stores. Nevertheless, the findings highlight key issues relevant to Dhaka and similar settings.

In summary, self-medication in urban Dhaka is common and driven by perceptions of mild illness, prior experience, and barriers to formal care. While most users are aware of potential harms and generally disapprove of the practice, many still resort to it regularly. These insights emphasize the need for multi-faceted interventions: enforcing prescription regulations, educating the public about safe medication use, and improving access to affordable medical services. Such measures could help curb irrational drug use and ensure that self-care is practiced responsibly.

## Conclusion

In this urban Bangladeshi study, self-medication for minor ailments was prevalent, particularly among young educated men. The practice was often irregular, with many users altering dosages or stopping treatment early. While nearly all obtained medicines from pharmacies, a significant proportion experienced side effect. Importantly, most participants recognized the risks of self-medication and viewed it as inappropriate, indicating a dissonance between behavior and belief. These results underscore the importance of enhancing community education on medication safety, reinforcing regulatory oversight of drug sales, and expanding healthcare access. Promoting responsible self-care through such measures may reduce the potential harms associated with unsupervised medication use.

## Data Availability

Data cannot be shared publicly because of ethical considerations. Data are available from the NIPSOM Institutional Data Access / Ethics Committee (contact via NIPSOM) for researchers who meet the criteria for access to confidential data.

## Acknowledgements

We thank all participants who participated in this study; and the community pharmacists for their cooperation. We also thank NIPSOM and Bangladesh Medical Research Council (BMRC) for their technical support respectively.

